# Development of the one-step qualitative RT-PCR assay to detect SARS-CoV-2 Omicron (B.1.1.529) variant in respiratory specimens

**DOI:** 10.1101/2022.01.04.22268772

**Authors:** Tung Phan, Stephanie Boes, Melissa McCullough, Jamie Gribschaw, Alan Wells

## Abstract

A new SARS-CoV-2 Omicron (B.1.1.529) Variant of Concern has been emerging worldwide. We are seeing an unprecedented surge in patients due to Omicron in this COVID-19 pandemic. A rapid and accurate molecular test that effectively differentiates Omicron from other SARS-CoV-2 variants would be important for both epidemiologic value and for directing variant-specific therapies such as monoclonal antibody infusions. In this study, we developed a real-time RT-PCR assay for the qualitative detection of Omicron from routine clinical specimens sampling the upper respiratory tract. The limit of detection of the SARS-CoV-2 Omicron variant RT-PCR assay was 2 copies/μl. Notably, the assay did not show any cross-reactivity with other SARS-CoV-2 variants including Delta (B.1.617.2). This SARS-CoV-2 Omicron variant RT-PCR laboratory-developed assay is sensitive and specific to detect Omicron in nasopharyngeal and nasal swab specimens.

## The study

SARS-CoV-2 is a new member of the large family of coronaviruses. Coronavirus disease (COVID-19) is an infectious disease caused by SARS-CoV-2 (1, 2). This virus was first known to infect people in 2019, and it has been reported to infect some animals (3, 4). SARS-CoV-2 can spread from person to person through droplets released when an infected person talks or coughs. In addition, there is strong evidence that SARS-CoV-2 can be transmitted by the airborne route (5, 6). Globally to date, SARS-CoV-2 has caused nearly 300 million COVID-19 cases and more than 5 million deaths (https://coronavirus.jhu.edu/map.html), making this pandemic one of the deadliest disease outbreaks in history.

Like other RNA viruses such as influenza viruses, SARS-CoV-2 has mutated over time, resulting in the high frequency of SNPs occurrence over the course of the COVID-19 pandemic (7, 8). Due to the rapid changes in the SARS-CoV-2 genome, the virus has been tracked in real-time by next-generation sequencing (NGS), and sequence data is shared openly in the most popular data-sharing platform GISAID to help analyzing how viral strains are spreading around the world (9). Given the significant genetic variation in the population of circulating SARS-CoV-2 strains, the FDA alerts clinical laboratories that the sensitivity of molecular, antigen and serology tests can be affected. The first case of the new SARS-CoV-2 Omicron Variant of Concern (VOC) from South Africa was reported to WHO on November 24^th^, 2021. The genomic analysis demonstrated that this Omicron VOC is classified into the Pangolin lineage B.1.1.529 and the Nextstrain clade 21K (https://www.cdc.gov/coronavirus/2019-ncov/science/science-briefs/scientific-brief-omicron-variant.html). The spike gene of Omicron is characterized by the large number of mutations (at least 30 amino acid substitutions), three small deletions and one small insertion. Of note, a half of amino acid substitutions are found to locate in the receptor binding domain, which can play an important role in ACE2 binding and antibody recognition. Importantly for the control and treatment of COVID-19, this is the same region that most vaccines present for immunity, and that therapeutic monoclonal antibodies target. Thus, these changes in the superspreading Omicron raises significant concerns about vaccines’ effectiveness and reduced efficacy of monoclonal antibodies treatments.

In the United States, Omicron was first detected on December 1^st^, 2021 in a traveler who returned from South Africa, and it is becoming the serious public health challenge (10). To rapidly detect Omicron in clinical specimens so as to direct therapies, we developed and evaluated the analytical and clinical performance characteristics of a SARS-CoV-2 Omicron variant RT-PCR assay for the qualitative detection of Omicron in the upper respiratory tract.

A total of 80 nasopharyngeal and nasal swabs in viral transport medium were used for the validation study. The specimens were previously SARS-CoV-2 tested by the Cepheid Xpert Xpress SARS-CoV-2/Flu/RSV test or the Hologic Aptima SARS-CoV-2 test. The specimens were extracted using the EasyMag (bioMérieux). The specimen input and elution output volumes were 200 μl and 70 μl, respectively. The SARS-CoV-2 Omicron variant RT-PCR assay included three separate RT-PCR reactions. Two primer/probe sets were designed based on the small deletion del69-70 and the small insertion ins214EPE seen in the Omicron spike gene (Fig. 1a). An additional primer/probe set to detect human RNase P gene (RP) in human specimens was also included in the assay (Fig. 1b). Primers and probes were purchased from Integrated DNA Technologies (IDT). Two primers and one probe in each set were pooled to generate a combined primer/probe mix in which the final concentration of primer and probe is 6.67 μM and 1.67 μM, respectively. For the RT-PCR reaction setup, 5 μl of extracted viral genome was added to 15 μl of a reagent mixture consisting of 5 μl of TaqPath 1-Step RT-qPCR Master Mix (ThermoFisher), 1.5 μl of combined primer/probe mix and 8.5 μl nuclease-free water. The total RT-PCR reaction volume was 20 μl. The RT step was performed at 25°C for 2 min and 50°C for 15 min, followed by incubation at 95°C for 2 min. The PCR step included 45 cycles of denaturation at 95°C for 3 sec, followed by annealing, extension, and data acquisition at 60°C for 30 sec on the ABI 7500 Fast Dx Real-Time PCR System (Applied Biosystems).

**Fig. 1.**
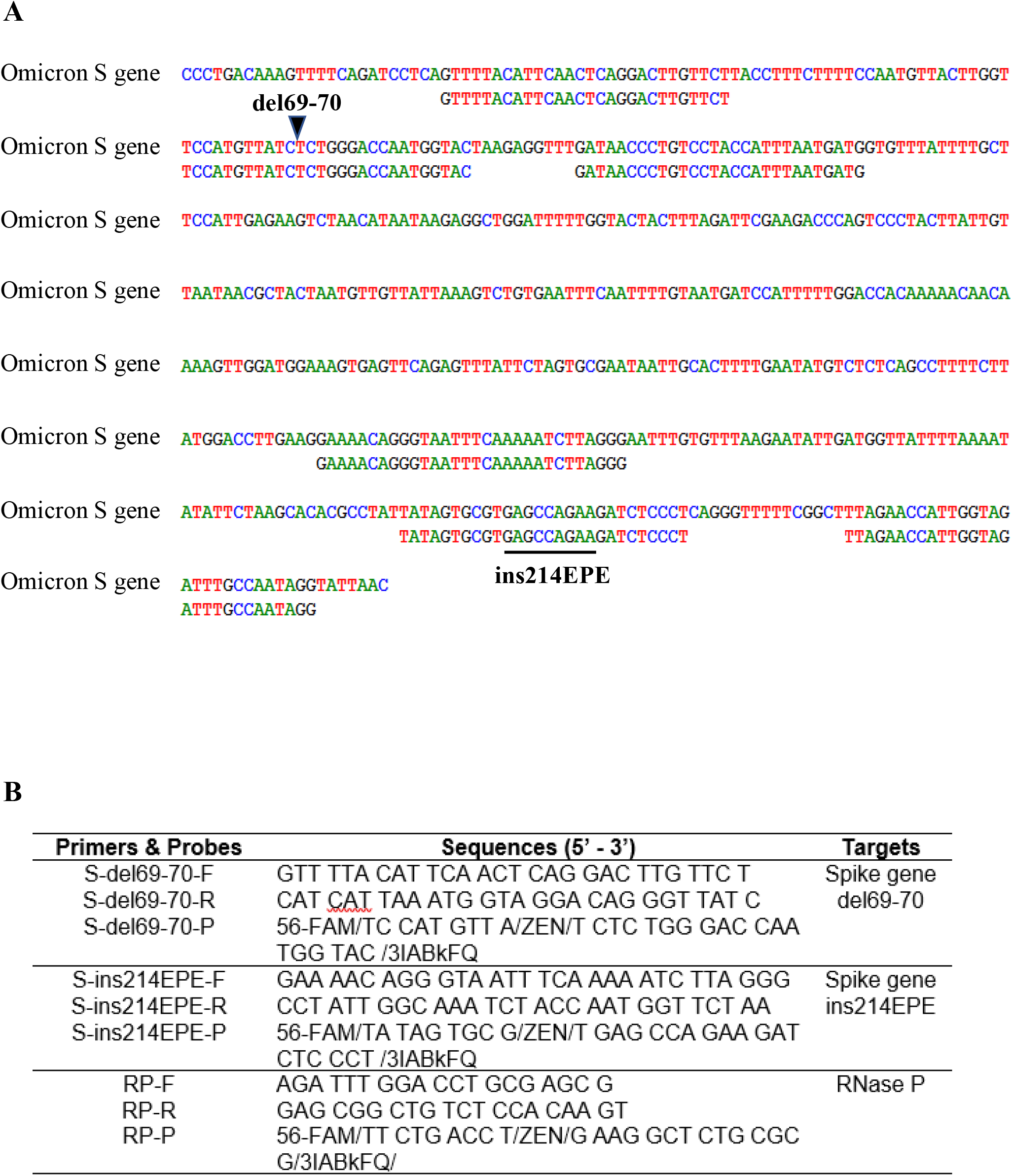
**A)** Alignment of the SARS-CoV-2 Omicron variant RT-PCR assay’s primers and probes to the spike gene sequence of SARS-CoV-2 Omicron (B.1.1.529) Variant of Concern. The small deletion del69-70 and the small insertion ins214EPE were shown. **B)** Names and sequences of the primers and probes used in the SARS-CoV-2 Omicron variant RT-PCR assay.

Since the quantitated Omicron variant is not available, we used the IDT gBlocks Omicron spike gene fragment dsDNA to determine the limit of detection (LOD) of the SARS-CoV-2 Omicron variant RT-PCR assay. The LOD was assessed by analyzing the IDT gBlocks Omicron spike gene fragment dsDNA simulated nasopharyngeal specimens with known titers ranging from 15 to 4,000 copies/ml. All specimen dilutions were prepared using a clinical negative nasopharyngeal matrix. The LOD titer is defined as the lowest concentration at which ≥95% of specimens tested generated positive calls. A minimum of 20 replicates were tested for the LOD verification. Table 1 showed that the positivity rate of 20 replicates observed was ≥95% at 1000 copies/ml for del69-70 (19/20, 95%) and at 2000 copies/ml for ins214EPE (20/20, 100%) targets. Since FDA requires claimed LOD based on the least sensitive target, the LOD of the SARS-CoV-2 Omicron variant RT-PCR assay was determined as 2000 copies/ml (or 2 copies/μl).

**Table 1:**
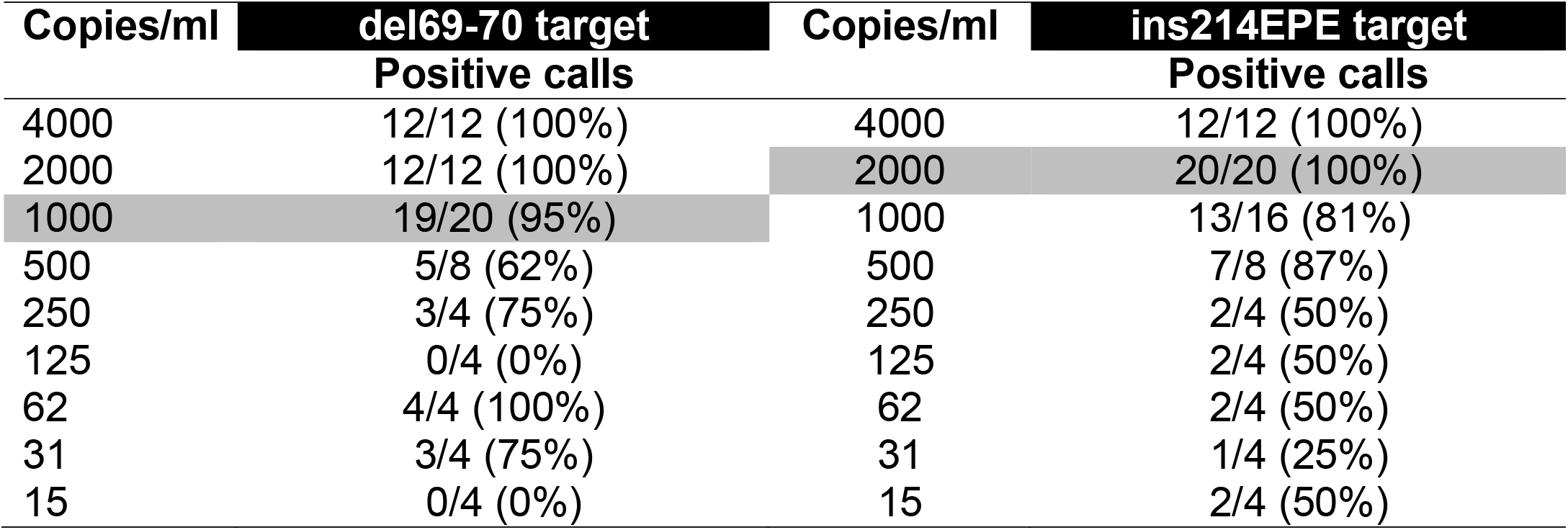
Limit of detection of the SARS-CoV-2 Omicron variant RT-PCR assay

To assess clinical performance characteristics of the SARS-CoV-2 Omicron variant RT-PCR assay, a total of 80 nasopharyngeal and nasal swabs (20 residual nasopharyngeal negatives, 20 residual nasal negatives, 20 simulated nasopharyngeal positives and 20 simulated nasal positives) were tested. The simulated specimens were made by spiking the IDT gBlocks Omicron spike gene fragment dsDNA in a clinical negative nasopharyngeal or nasal matrix to generate 10 different levels from 2X LOD to 1024X LOD. We did not see any false positives or false negatives when tested the negative or positive specimens, respectively. The clinical sensitivity was 100%, the clinical specificity was 100%, and the agreement was 100% as shown in Table 2. The SARS-CoV-2 Omicron variant RT-PCR assay’s cross-reactivity was evaluated by testing the ability of the assay to exclusively identify Omicron with no cross-reactivity to organisms that are closely related or cause similar clinical symptoms or may be present in nasopharyngeal or nasal specimens. AccuPlex™ SARS-CoV-2 Variant Panels were purchased from SeraCare (Life Sciences, Inc) and tested by the SARS-CoV-2 Omicron variant RT-PCR assay. The assay did not amplify the Wuhan wild type and other variants including B.1.351 (Beta), B.1.617.1 (Kappa), P.1 (Gamma), B.1.617.2 (Delta) and AY.1 (Delta Plus). While the del69-70 target was detected in the variant B.1.1.7 (Alpha) by the SARS-CoV-2 Omicron variant RT-PCR assay, the ins214EPE target was undetected. This observation is expected because del69-70 is seen in both B.1.1.7 (Alpha) and Omicron. No cross-reactivity with other respiratory viruses including influenza A(H3) virus, influenza B virus, respiratory syncytial virus A & B, common coronaviruses (OC43, NL63 and 229E), and bacteria including *Klebsiella pneumoniae* and *Streptococcus pneumoniae* (ZeptoMetrix) was seen in Table 3.

**Table 2.** Sensitivity, specificity & agreement of the SARS-CoV-2 Omicron variant RT-PCR assay

| **Omicron variant assay** | **True positive** | **True negative** |
| --- | --- | --- |
| Positive | 40 | 0 |
| Negative | 0 | 40 |
| Sensitivity | $(40+0) / (40+0)=100\%$ | |
| Specificity | $(0+40) / (0+40)=100\%$ | |
| Agreement | $(40+40) / (40+40)=100\%$ | |

**Table 3.** Cross-reactivity of the SARS-CoV-2 Omicron variant RT-PCR assay. The experiment was assayed in triplicate.

| **Organisms** | **Observed result** |  | **Omicron variant assay's interpretation** |
| --- | --- | --- | --- |
|  | **del69-70 target** | **ins214EPE target** |  |
| Wuhan wild type | Undetected | Undetected | Negative |
| B.1.1.7 (Alpha) | Detected | Undetected | Negative |
| B.1.351 (Beta) | Undetected | Undetected | Negative |
| P.1 (Gamma) | Undetected | Undetected | Negative |
| B.1.617.1 (Kappa) | Undetected | Undetected | Negative |
| B.1.617.2 (Delta) | Undetected | Undetected | Negative |
| AY.1 (Delta Plus) | Undetected | Undetected | Negative |
| Coronavirus 229E | Undetected | Undetected | Negative |
| Coronavirus NL63 | Undetected | Undetected | Negative |
| Coronavirus OC43 | Undetected | Undetected | Negative |
| Influenza A(H3) virus | Undetected | Undetected | Negative |
| Influenza B virus | Undetected | Undetected | Negative |
| RSV A | Undetected | Undetected | Negative |
| RSV B | Undetected | Undetected | Negative |
| K. pneumoniae | Undetected | Undetected | Negative |
| S. pneumoniae | Undetected | Undetected | Negative |

The Omicron variant has turned out to be more contagious than any earlier coronavirus strains. The early research suggested that Omicron mutations enhance infectivity and reduce the neutralization of antibodies formed by prior infection or vaccination (11). Thus, rapid identification would help those patients at high risk of succumbing to COVID 19. While there are a variety of platforms currently being used to identify SARS-CoV-2, commercial assays specific for Omicron detection are not available. While the S-gene target failure (SGTF)-based method is widely used to flag potential Omicron cases, it is not accurate since SGTF is not unique to Omicron. The 69-70 deletion that reduces S-gene target amplification in some molecular assays is also found in Alpha variant along with some other variants (12). Further, negative identification is not as reliable a technique as positive identification.

At present, NGS is the best option to genotype SARS-CoV-2 (13); however, this approach is not fast enough and lack adequate throughput for the current surge in cases. In addition, only a few clinical laboratories can perform complicated NGS technology assays. Therefore, developing a rapid and accurate molecular test effectively differentiates Omicron from other SARS-CoV-2 variants in clinical specimens is very important. Here, we successfully developed the new one-step qualitative RT-PCR assay to detect Omicron in respiratory specimens. While our assay was evaluated on the ABI 7500 Fast Dx Real-Time PCR System (Applied Biosystems), clinical laboratories can validate it on other available Real-Time PCR systems. Our assay was designed to target the small deletion del69-70 and the small insertion ins214EPE seen in the Omicron spike gene, and human RNase P gene as an internal control. A specimen is considered positive only if two targets (del69-70 and ins214EPE) are detected. During the validation, precision analysis was also performed by testing two negative and two positive specimens, which were run by three different technologist, and all of them (100%) yielded expected results. In the carry-over contamination study, we ran the negative nasopharyngeal specimens next to the positive nasopharyngeal specimens with a high titer (10^6^ copies/ml). All the negative specimens remained negative, suggesting that the carry-over contamination did not happen.

## Data Availability

All data produced in the present work are contained in the manuscript

## Acknowledgements

We thank the UPMC Clinical Microbiology Laboratory for testing the specimens and performing the evaluation.

## Funding

The study was internally funded by the UPMC Clinical Laboratories as part of a Quality Improvement initiative. No funding was obtained from any commercial sources. The funders did not have input into study design, analysis, nor generation of this communication.

## Author contributions

TP and AW: designed the study and wrote the manuscript; SB, JG and MM: performed and managed the testing.

## Conflicts of interest

The authors declare no competing financial interests.

## Ethical approval

All testing was performed as apart of routine clinical care and performed according to CLIA ‘88 regulations by appropriate personnel. The entire study was deemed to be a Quality Improvement initiative by the UPMC IRB and approved by the UPMC QI Review Board.

## Notes

### Competing Interest Statement

The authors have declared no competing interest.

### Author Declarations

The entire study was deemed to be a Quality Improvement initiative by the UPMC IRB and approved by the UPMC QI Review Board.

